# Influence of setting and diagnostic algorithm on disease severity among people diagnosed with symptomatic and asymptomatic tuberculosis in South Africa

**DOI:** 10.64898/2026.07.10.26357755

**Authors:** Nicky McCreesh, Indira Govender, Mareca Sithole, Emily B. Wong, Innocentia Buthelezi, Willem A. Hanekom, Gregory Ording-Jespersen, Mark J. Siedner, Alison D. Grant, Palwasha Y. Khan

**Author notes:** Corresponding author. Nicky McCreesh, London School of Hygiene & Tropical Medicine, London, WC1E 7HT, UK. Joint senior author (Alison D. Grant, Palwasha Y. Khan).

## Abstract

**Background:** There is growing interest in tuberculosis (TB) community screening and detection of asymptomatic TB (aTB). We explored how setting and screening approach influence the relationship between reported symptoms and underlying disease severity and infectiousness.

**Methods:** We compared markers of TB severity and infectiousness (computer-aided detection [CAD, CAD4TBv5] scores derived from chest radiographs and trace vs exceeding trace Xpert MTB/RIF Ultra results) among people diagnosed with aTB or symptomatic TB (sTB) through a community survey, and those diagnosed with aTB or sTB in clinics in the same South African community. We evaluated how variation in screening algorithms influence the relative severity of community-diagnosed aTB vs sTB, and estimated the TB prevalence and severity among people not eligible for testing in the survey (CAD score <25 and no reported symptoms).

**Results:** People with clinic-diagnosed sTB had higher CAD scores and a greater proportion of Xpert results exceeding trace than those with community-diagnosed sTB, whereas differences between community-diagnosed aTB and sTB were minimal. Under a hypothetical community universal Xpert testing strategy, people detected with sTB may have more severe disease on average than people detected with aTB. In contrast, restricting testing to people with CAD scores ≥50 and/or reported symptoms would have resulted in higher CAD scores among those diagnosed with aTB than sTB.

**Conclusions:** Screening algorithm design and context influence the spectrum of TB disease detected. Community screening approaches where testing is based on symptoms and/or a threshold CAD score may identify aTB that is on average more severe than sTB.

**Summary:** Our findings suggest reported symptoms are not a reliable indicator of TB disease severity, and that the context in which someone is investigated for TB – self-presentation to a clinic vs active community screening – may be more informative.

## Background

TB care and prevention in low- and middle-income settings has traditionally relied largely on passive case finding, requiring people to recognise symptoms and seek clinical care. However, with global TB incidence declining by only ∼1% per year[1], it is increasingly clear that passive detection alone is insufficient to achieve substantial reductions in population-level burden. This has led to a renewed interest in active case finding (ACF), where systematic screening is conducted in defined high-risk populations or communities. Concurrently, national prevalence surveys have revealed a substantial reservoir of TB in people who do not report symptoms (asymptomatic TB (aTB))[2], highlighting the potential importance of TB missed by symptom-based screening. Advances in digital radiography and computer-aided detection (CAD) algorithms have further accelerated this shift, enabling the rapid expansion of radiologically driven ACF, and making the detection of aTB increasingly feasible[3, 4].

This evolving diagnostic landscape raises fundamental epidemiological questions regarding the infectiousness of aTB and symptomatic TB (sTB) detected through community screening versus clinic-based diagnosis, their different clinical trajectories and therapeutic needs, and the implications of these distributions for forecasting future disease burden[5]. A clearer understanding is needed of how the setting and screening algorithm used in community ACF shape the spectrum and severity of disease identified.

The aim of this paper is to compare indicators of disease severity and infectiousness (CAD scores and proportion with Xpert output greater than trace) between people diagnosed with aTB and sTB in clinics or during a community, using data from two studies conducted in the same South African community. We further explore how the choice of screening algorithms and CAD thresholds might influence the severity profile of aTB and sTB detected through community screening.

## Methods

### Setting

This descriptive epidemiological study uses data from two cross-sectional studies conducted in the area of the Africa Health Research Institute (AHRI) health and demographic surveillance system (HDSS) in uMkhanyakude district in KwaZulu-Natal, South Africa, described below.

### Studies

#### Community survey

The community survey was conducted in the AHRI HDSS in 2018 – 2019, as part of a multimorbidity study (Vukuzazi, see [6] for full details). 18,041 people aged at least 15 years were recruited to the community survey, which included HIV testing and TB screening (symptom screen and chest x-ray (CXR) read using CAD4TB version 5). Individuals who reported any ‘WHO 4-symptom screen’ symptoms[7] (cough of any duration, fever, night sweats, and/or weight loss), had a CAD score above the threshold (60 for the first 7.8% of participants, reduced to 25 for the remaining 92.2%), or had abnormal lung fields as determined by a radiologist were eligible to have sputum samples collected and tested for *Mycobacterium tuberculosis* with an Xpert MTB/RIF Ultra test (Cepheid, Sunnyvale, CA, USA) and liquid mycobacterial culture (BACTEC MGIT 960 System [Becton Dickinson, Berkshire, UK]). Individuals were also eligible for sputum testing if they could not undergo a CXR (e.g. due to pregnancy), or if testing was indicated for other reasons (e.g. other reported symptoms).

Data from people who were screened in the community survey before the CAD threshold testing threshold was changed to 25, people who reported that they were currently receiving TB treatment at the time of enrolment, and people who did not complete the symptom screening were excluded from these analyses. For details, see supporting figure S1.

In the community survey, people with TB were defined as individuals with a positive result on Xpert (trace included) and/or culture. As culture is not typically conducted as part of community active case finding interventions, we defined people with TB as people with a positive result on Xpert (trace included), excluding people who were positive on culture only from our definition of ACF-detectable TB in the main analysis. Results of sensitivity analyses including people with Xpert undetected culture positive TB are shown in the supplementary information (Table S1 and Figures S2-S4).

Ethical approval for the study was obtained from the Ethics Committees of the University of KwaZulu-Natal (BE560/17), London School of Hygiene & Tropical Medicine (#14722), the Partners Institutional Review Board (2018P001802), and from the University of Alabama at Birmingham (#300007237). Written informed consent was obtained from all participants, and participant information anonymised.

#### Clinic-based recruitment

The clinic-based study recruited people aged 15+ years starting treatment for bacteriologically-confirmed pulmonary tuberculosis in clinics serving the AHRI HDSS between February 2022 and December 2023 (see Khan *et al* 2026[8] for full details). Data were not collected on whether people were detected through active vs passive screening. Based on our knowledge of how screening was being conducted in the clinics at the time however, it is probable that the majority of people not on ART (HIV negative or PWH not on ART) self-presented with symptoms, whereas a greater proportion of people on ART were likely to have been diagnosed through active screening. People were excluded if they had a history of TB treatment within the past two years, or if they did not live with any children aged 2 – 14 years. At enrolment, participants were asked about TB symptoms and had a digital CXR taken. To enable comparisons to be made with people diagnosed with TB as part of the community survey, we had the CXRs read by CAD4TB version 5.

The study was approved by the Biomedical Research Ethics Committee of the University of KwaZulu-Natal (BF472/19) and the Ethics Committee of the London School of Hygiene & Tropical Medicine, UK (17728-6). Written informed consent was obtained from all participants, and participant information anonymised.

### Analysis

#### Asymptomatic and symptomatic TB case definition

In both studies, people were considered to have TB if they had a positive Xpert Ultra result (trace included). In the clinic-based study, this included 1) people with a positive Xpert Ultra result (trace included) on a test ordered programmatically by the clinic, and 2) people positive on a programmatic smear microscopy test and negative on a programmatic Xpert Ultra test, who had a positive Xpert Ultra result on a subsequent research sputum specimen.

In the community screening study, the division into trace or greater than trace Xpert result was based on the results of the single Xpert Ultra test conducted during screening. In the clinic-based study, it was based on the programmatic Xpert Ultra result if the programmatic Xpert was positive. Individuals with a negative programmatic Xpert result and positive programmatic smear were considered to be trace or greater than trace based on the lowest of two subsequent Xpert Ultra tests conducted as part of the research study.

We considered that someone with TB had sTB if they reported one or more ‘WHO 4-symptom screen’ symptoms[7] (cough of any duration, night sweats, fever, or weight loss), and aTB if they reported none of the four symptoms at the point of study enrolment.

#### Comparison of aTB and sTB diagnosed in clinics vs the community survey

As an indicator of disease severity and infectiousness, we compared CAD scores and proportions of people with Xpert greater than trace between people diagnosed with aTB and sTB in the clinics vs in the community survey, overall and by HIV/ART status.

#### Effect of choice of screening algorithms and CAD threshold

To investigate how the choice of screening algorithms and CAD thresholds might influence the severity of aTB and sTB detected through community screening, we calculated the median CAD scores and the proportion of people with Xpert results above trace for a number of screening and testing scenarios. Firstly, if CXR screening and symptom screening were conducted in parallel, with the CAD4TB v5 threshold set to 25, 30, 40, 50, 60, or 70. Secondly, if only CXR screening was used, with the CAD threshold set to the same range of values. In these analyses, we assumed that the testing criteria would be applied strictly, with people only tested if eligible under the screening algorithm. Finally, if no screening step was used i.e. everyone was eligible for Xpert testing regardless of symptoms or CAD score. Participants with no CAD score available were excluded from these analyses.

As the majority of people with CAD4TB score <25 who did not report symptoms were not tested, we estimated the proportion who would have been diagnosed with TB, and their CAD scores and Xpert result, had they been eligible for testing. This was done using bootstrapping, assuming that the prevalence and severity of TB in people with CAD score <25 was the same for people who did not report symptoms as for people who did. Additional details are given in the supporting material text and Table S2.

## Results

### Comparison of aTB and sTB diagnosed in clinics vs the community survey

A total of 134 people from the community survey were included in our analyses of people considered to have TB, 115 (86%) of who had asymptomatic TB (Table 1). Two people who reported no WHO 4-symptom screen questions and had CAD score <25 received a positive Xpert result. The median CAD score among respondents included in our analyses was 64 (IQR 44 – 78) and 73 (55%) had an Xpert result greater than trace.

**Table 1.**
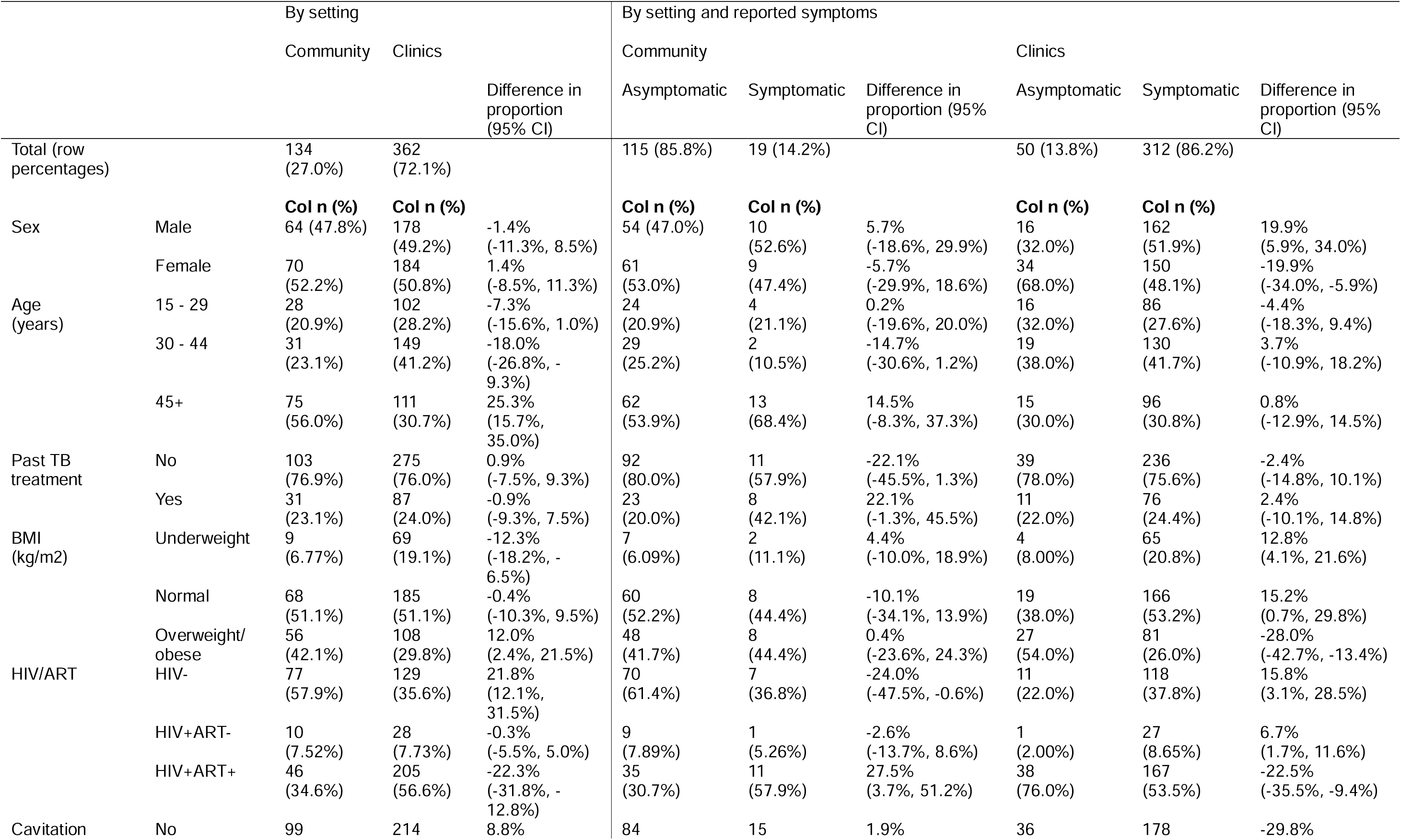
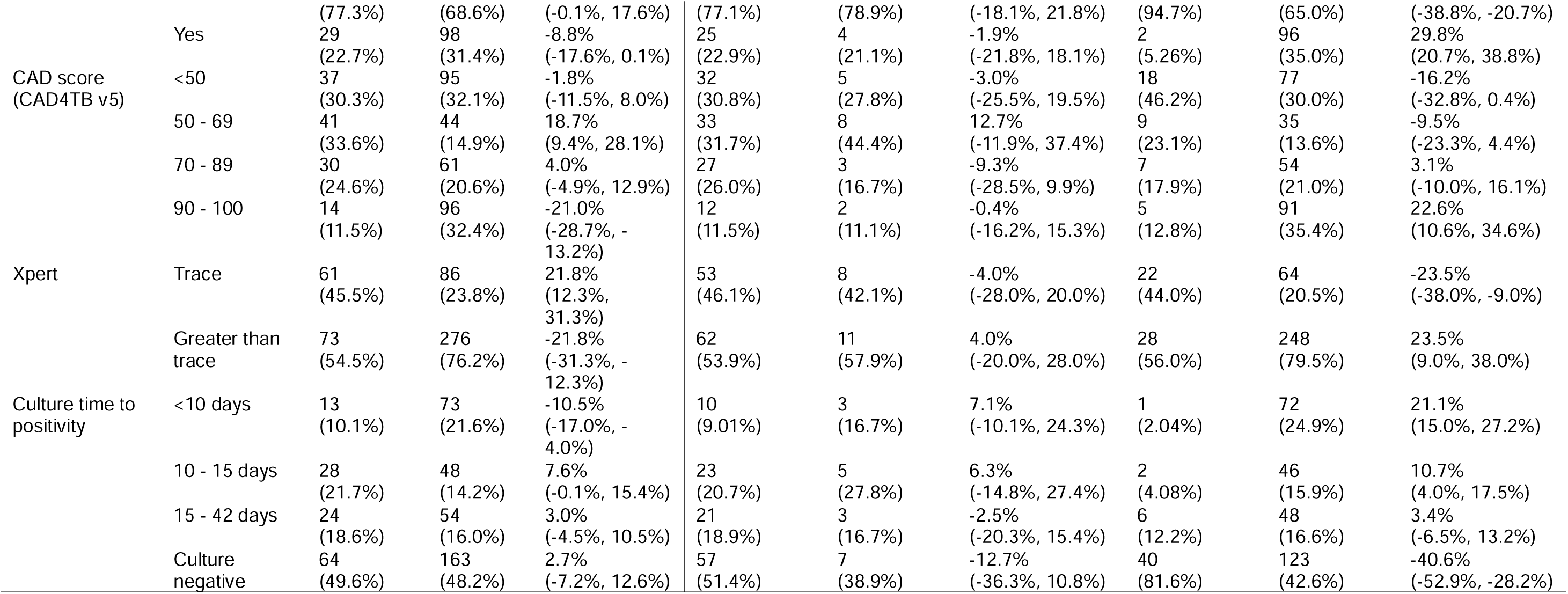
Demographic and clinical characteristics of people diagnosed with aTB or sTB in the clinic and community surveys. In people diagnosed with TB as part of the community survey, data were missing on BMI for one person, HIV/ART for one person, cavitation and CAD score for six people, and time to culture positivity for five people. In people diagnosed with TB in clinics, data were missing on cavitation from 50 people, CAD score from 49 people and time to culture positivity for 24 people. Confidence intervals for differences in proportions (risk difference) are estimated using the Score method.

A total of 362 people from the clinic-based study were included, 50 (14%) of who had asymptomatic TB. 360 participants had a positive result on a programmatic Xpert test. The remaining two had a positive result on a programmatic smear microscopy test and negative result on a programmatic Xpert Ultra test, followed by two positive “high” results on subsequent research sputum specimens. The median CAD score was 81 (IQR 42 – 95) and 276 (76%) had an Xpert result greater than trace.

Compared to people diagnosed in clinics, people diagnosed with TB in the community survey were older, and less likely to be underweight, HIV positive, have a CAD score > 90, or have a time to culture positivity <10 days (Table 1). People diagnosed in the community with sTB compared to aTB were more likely to have reported past TB treatment. The proportion of people diagnosed with TB who reported symptoms was substantially higher in clinics (312/362, 86.2%) than in the community (19/134, 14.2%).

People with sTB in the clinic study had higher CAD scores than people with sTB in the community survey, overall (median CAD score 83 clinic, 63 community, p=0.1), and when limited to HIV-negative people (median CAD score 91 clinic, 64 community, p=0.04) (Figure 1). People with aTB in the clinic study had lower CAD scores than people with aTB in the community survey, overall (median CAD score 53 clinic, 63.5 community, p=0.02), and when limited to PWH on ART (median CAD score 44.5 clinic, 64 community, p=0.02). In the clinic study, people with sTB had higher CAD scores than people with aTB, overall and by HIV/ART status (overall: sTB 83, aTB 53, p<0.0001; HIV-negative: sTB 91, aTB 58.5, p=0.02; PWH on ART: sTB 63, aTB 44.5, p=0.005). In the community survey, there was minimal difference in CAD scores between people with sTB and people with aTB, overall and by HIV/ART status.

**Figure 1.**
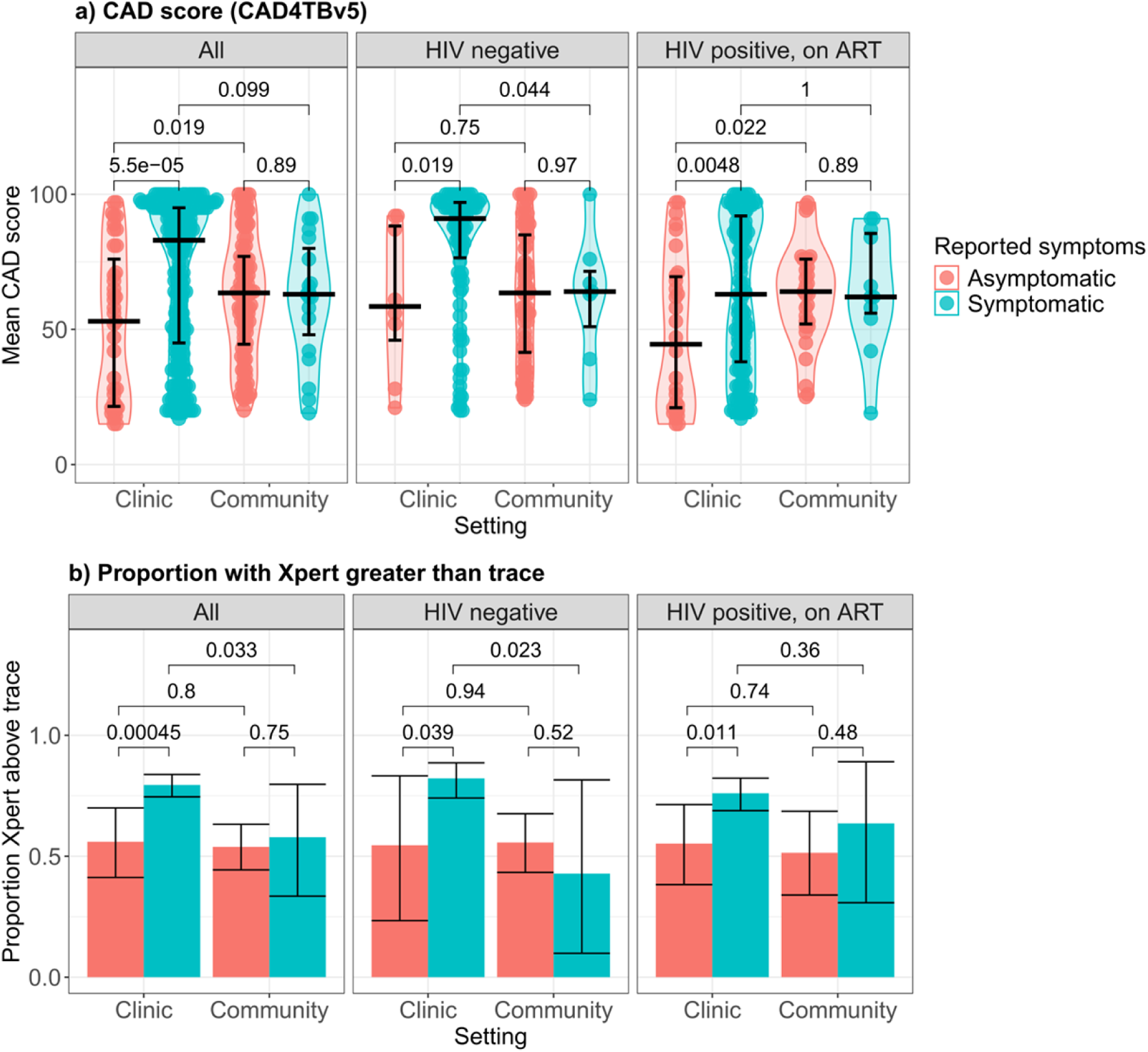
Comparison of CAD4TB v5 scores (top row) and proportion of people who were Xpert trace or above (bottom row) in people diagnosed with TB, by setting (clinic vs community), symptoms, and HIV/ART status. Results for people with HIV who were not on ART are not shown due to very small numbers (only one person diagnosed with sTB in the community, one only one with aTB in a clinic), however are shown in supporting figure S2. Top row: horizontal lines show median and interquartile range, and the numbers on the plots are p-values, calculated using a Mann-Whitney U-Test. Bottom row: error bars show 95% confidence intervals, and the numbers on the plots are p-values, calculated using a Wald test. Alt text: Six plots showing the distribution of CAD scores and the proportion of people with an Xpert result greater than trace, by HIV/ART status, and by diagnostic setting and reported symptoms

The proportion of people with sTB with Xpert result greater than trace was higher for people in the clinic study than the community survey, overall (clinic 79%, community 58%, p=0.03), and when limited to HIV-negative people (clinic 82%, community 43%, p=0.02) (Figure 1). There was minimal difference between the clinic study and community survey in the proportion of people with aTB with Xpert result greater than trace, overall and by HIV/ART status. The proportion of people in the clinic study with Xpert result greater than trace was higher for people with sTB than aTB, overall (sTB 79%, aTB 56%, p<0.001), when limited to HIV-negative people (sTB 82%, aTB 55%, p=0.04), and when limited to PWH on ART (sTB 76%, aTB 55%, p=0.01). There was minimal difference between people with aTB vs sTB in the community survey in the proportion of people with Xpert result greater than trace, overall and by HIV/ART status.

### Effect of choice of screening algorithms and CAD threshold

If no screening step had been used in the community survey (i.e. everyone had been asked to give a sputum sample), we estimated that a mean of 33 (95% CI 0 – 83) people would have been diagnosed with aTB among those with a CAD score <25, in addition to the 107 people with aTB and CAD score ≥25 who were identified.

Figure 2 shows how the average relative severity of aTB and sTB detected during community screening may vary with the choice of CAD threshold, assuming that all people who report symptoms and/or have a CXR scoring above the threshold are eligible for testing. As expected, at higher CAD thresholds the median CAD score of aTB diagnosed through screening increases, and at thresholds above 40 people diagnosed with aTB would have had significantly higher CAD scores than people diagnosed with sTB. In contrast, we estimate that if all people had been eligible for testing, regardless of symptoms or CAD score, then median CAD scores and the proportion with Xpert greater than trace would have been lower in people diagnosed with aTB compared to sTB, although the differences were not significant.

**Figure 2.**
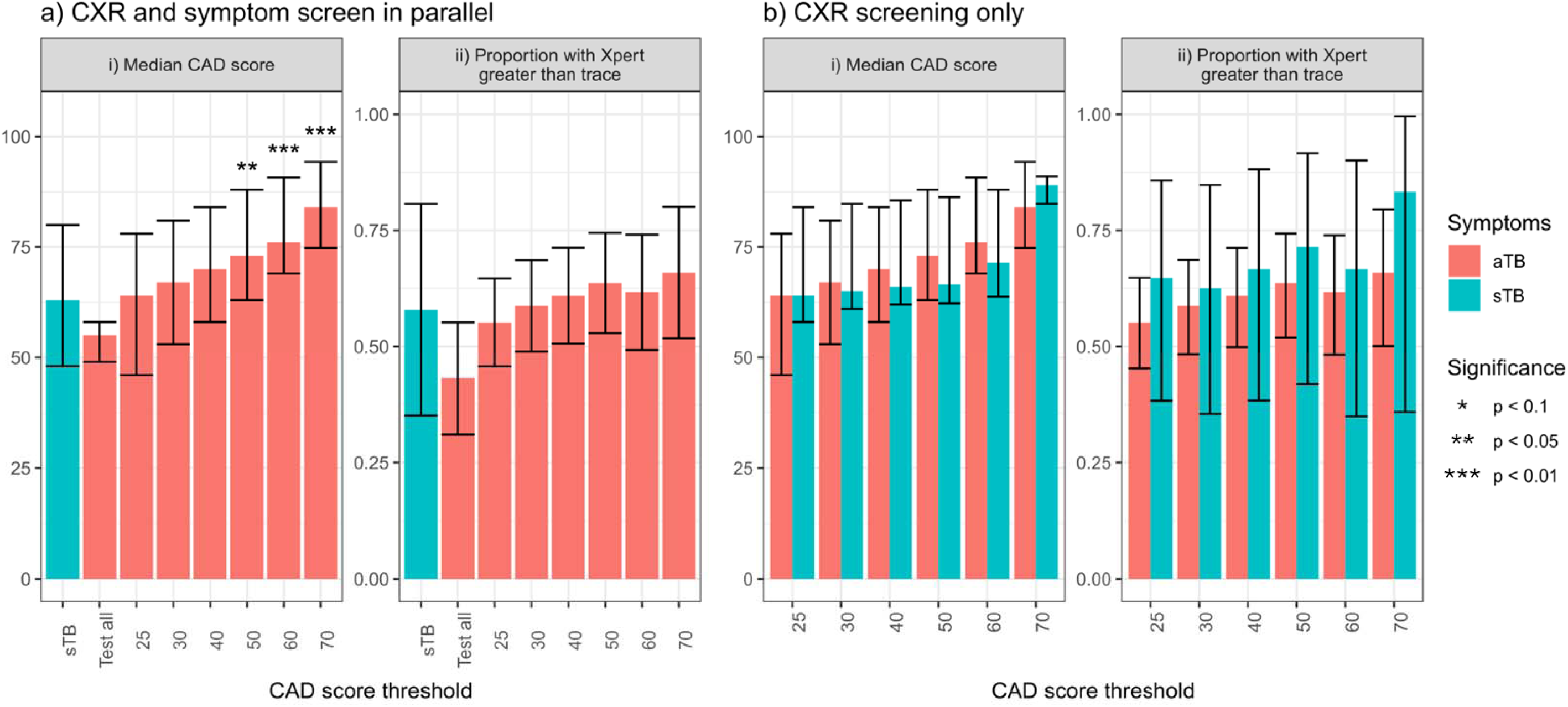
Effect of choice of CAD cut-off used in screening on relative severity of aTB and sTB diagnosed in the community. In a), it assumed that all people who report symptoms will be eligible for testing, but people who do not report symptoms will be eligible for testing only if they have a CAD score above the threshold. In b) it is assumed that people will be eligible for testing only if they have a CAD score above the threshold, regardless of reported symptoms. The bars Plots a – d: Asterisks indicate p-values for differences between aTB and sTB, where p<0.1. Where no asterisk is shown, p≥0.1. i) The bars show the median, and the error bars the inter-quartile range. p-values calculated using a Mann-Whitney U-Test. ii) The bars show the mean, and the error bars the 95% confidence interval. p-values calculated using a Wald test Alt text: Four bar charts showing how median CAD scores and proportion with an Xpert result greater than trace vary with screening approach and CAD threshold

## Discussion

Our findings suggest reported symptoms (based on the WHO four symptom screen[7]) are not a reliable indicator of TB disease severity, and that the context in which someone is investigated for TB – in a clinic vs active community screening – may be more informative. Comparing people diagnosed with sTB vs aTB in the community survey, there is little evidence that sTB represents a more severe or more infectious state on average, with no difference found between CAD scores or the proportion with Xpert above trace among people diagnosed with sTB vs aTB. In contrast to this, we find evidence that sTB identified among people self-presenting in clinics may be more severe on average than aTB or sTB diagnosed in the community, and more severe than aTB diagnosed in clinics. In all four groups, there was substantial heterogeneity between individuals, e.g. with CAD scores ranging from 20 or lower to 97 or higher.

Our results also highlight the importance of screening algorithm design on the spectrum of disease severity identified in people diagnosed with aTB vs sTB. We show that if universal testing is conducted, with no screening step, then sTB may be more severe on average than aTB (although the differences were not significant in our study). At the other extreme, with parallel symptom and CAD screening, CAD scores may be substantially higher in people diagnosed with aTB than people diagnosed with sTB, reflecting the diagnosis of below CAD threshold TB in people reporting symptoms. Figure 3 shows a conceptual diagram demonstrating how these differences can occur.

**Figure 3.**
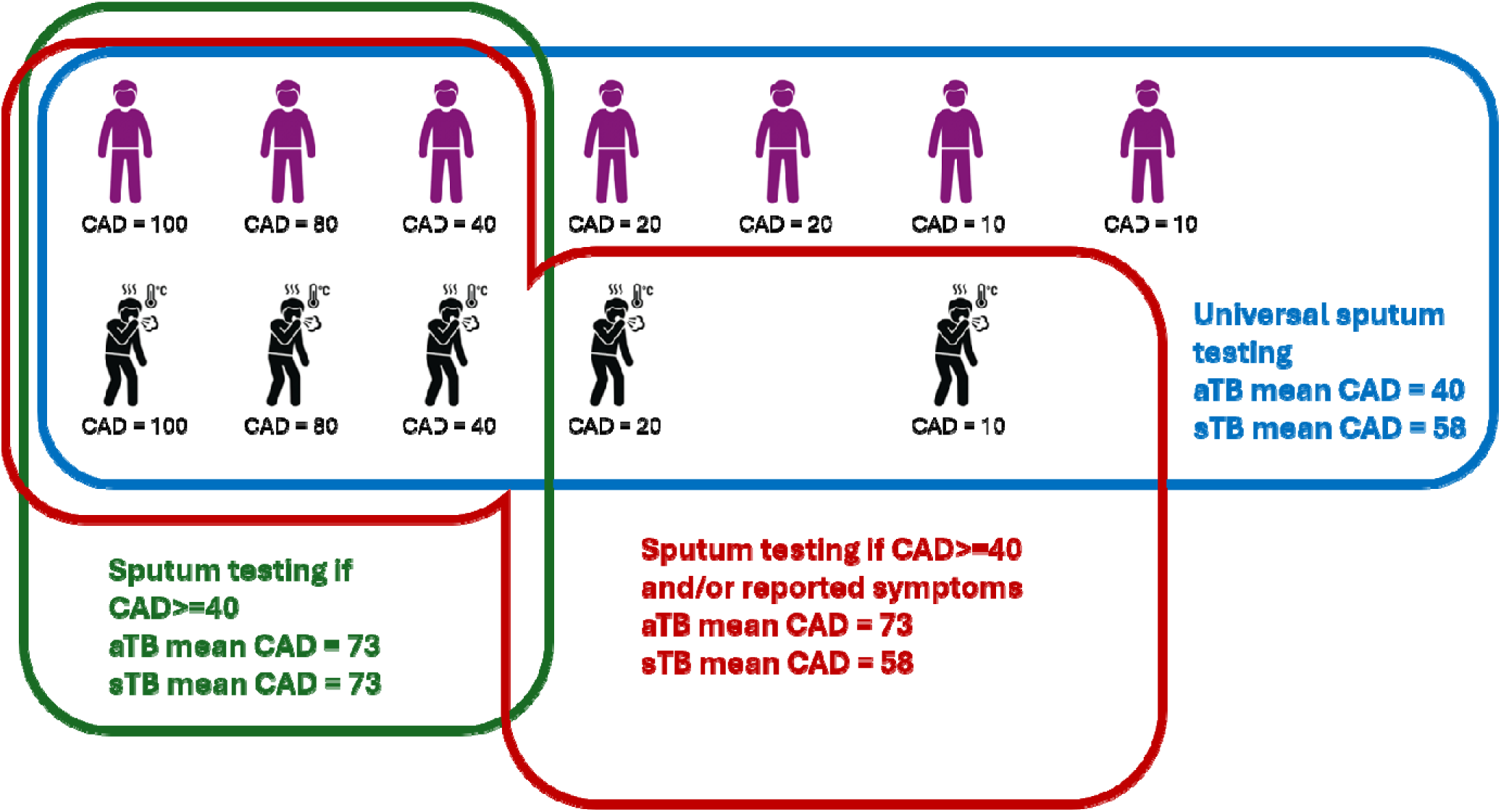
Conceptual diagram demonstrating how the choice of screening algorithm can affect the relative severity of asymptomatic TB (aTB) vs symptomatic TB (sTB) diagnosed during community screening. Purple figures indicate people with aTB, and black figures people with sTB. The text underneath each figure shows their CAD score. The blue, red, and green shapes indicate the people who would be detected through three different screening algorithms, and the mean CAD scores in people detected with aTB and sTB Alt text: Icons of seven sick and five healthy individuals with their CAD scores underneath. Three shapes encircle the individuals who would be detected through three different screening algorithms, and text indicates the mean CAD scores in people diagnosed with aTB vs sTB through each algorithm

These results have major implications for how we think about the natural history of TB, and how we group and classify the spectrum of disease. We provide evidence that sTB should not be considered a homogeneous state. As a result, findings from people self-presenting to clinics with sTB (e.g. data on diagnostic test performance or the requirements for effective drug regimens) cannot necessarily be generalised to the full spectrum of sTB in the community. Conversely, given the substantial overlap in disease severity between people diagnosed with aTB and sTB in community settings, and dependence of severity on the screening algorithm, it is unsurprising that studies aiming to, for instance, compare the infectiousness of community diagnosed aTB vs sTB have not found consistent evidence for a difference[9–11].

Our results differed between HIV-negative people and PWH on ART. Among HIV-negative people, disease severity was substantially higher in those diagnosed with sTB in clinics compared with the community, whereas there was no corresponding difference in severity between clinic- vs. community-identified aTB. This likely reflects the fact that (outside of ART care) testing in clinics was predominantly passive, requiring symptom-driven care seeking. For PWH on ART, the pattern was reversed – there was no evidence for any difference in disease severity between people diagnosed with sTB in clinics vs the community, but CAD scores were lower in people diagnosed with aTB in clinics. The absence of differences for sTB in this group may reflect routine active screening among ART clinic attendees, making clinic- and community-diagnosed TB more comparable. Lower CAD scores among clinic-vs. community-diagnosed aTB likely reflects community survey testing criteria, with the majority of asymptomatic people only tested if their CAD score was ≥25.

Our finding that sTB diagnosed in clinics may be more severe on average than sTB diagnosed in the community is in line with the results of previous studies. Kendall *et al.* found that 77% of people diagnosed with sTB in clinics had serum C-reactive protein (CRP) levels of >10mg/L, compared to only 34% of people diagnosed with sTB as part of community screening[12]. Mendelsohn *et al* found substantially higher median CRP level in people diagnosed with sTB in clinics than sTB diagnosed in household contacts (17.8mg/dL vs 4.3mg/dL)[13]. Mtshali *et al* compared IL-6 and CRP levels between people diagnosed with sTB during the same community survey that we used in this paper and a different sample of 30 people diagnosed with sTB in clinics, finding substantially higher levels in people diagnosed in clinics (IL-6 11.3 vs 0.79pg/ml, CRP 88mg/L vs 7mg/L)[14].

Comparing sTB and aTB diagnosed in the community, the same studies have found more mixed results. Kendall *et al*.’s data show that 82% of people diagnosed with Xpert positive aTB in the community had trace Xpert results, compared to only 49% of people diagnosed with Xpert positive sTB[12]. Mendelsohn *et al* found higher median CRP levels in household contacts with sTB compared to aTB, although the difference was not significant (4.3 vs 1.7, p=0.3)[13]. Mtshali *et al* found no evidence for any difference in IL-6 and CRP levels in people diagnosed with sTB vs aTB during the community survey (IL-6 0.79 vs 0.79pg/ml, CRP 7mg/L vs 5mg/L)[14].

One potential contributor to the difference may be that both Kendall *et al* and Mendelsohn *et* al tested everyone, regardless of symptoms or CXR readings. Our extrapolated results suggest that, had everyone been tested in the community survey, CAD scores and proportions with Xpert results greater than trace would have been lower in people diagnosed with aTB compared to sTB, in line with the results of other studies. A second potential contributor is that the definition of TB symptoms varied between the studies. This study, Mtshali *et al*, and Mendelsohn *et al*, used a broader definition of symptoms (cough of any duration, fever, weight loss, and/or night sweats, with the addition of fatigue, pleuritic chest pain, and haemoptysis by Mendelsohn *et al*). Kendall *et al*. used a more restrictive definition (cough ≥**2 weeks**, fever, night sweats, and/or weight loss), which could plausibly result in greater differences in disease severity between people diagnosed with aTB vs sTB in the community.

There are a number of limitations of our study. In the community survey, testing was restricted to people at higher risk of TB – predominantly people who reported symptoms and/or had CAD score ≥25. Estimates of the potential severity of aTB vs sTB that would be detected by universal testing in the community are therefore extrapolated. Our estimates of the potential severity of aTB under universal testing should therefore be interpreted as giving a qualitative indication of the effect of expanding testing, rather than as accurate numerical estimates. Differences between how the different tests were conducted in the clinic and community survey studies (e.g. length of time between sputum collection and testing) may have biased the comparisons between TB detected in clinics vs the community. Participants in the community survey were asked about symptoms at the point of screening, whereas participants in the clinic study were asked about symptoms at study enrolment, a few days after giving their programmatic sputum sample – potentially increasing symptom reporting[15], particularly among people with milder symptoms. Numbers of people diagnosed with aTB in clinics or sTB in the community survey were relatively low, reducing the power of the study. Data were not collected on whether people diagnosed with sTB in clinics were detected through passive or active case finding, preventing us from stratifying the analysis by reason for testing. Finally, while not a limitation of this study *per se*, it should be noted that symptom reporting is highly subjective, and may vary substantially between different settings[16]. It is therefore unclear the extent to which the results of this study, and other studies comparing sTB and aTB, can be generalised outside of the immediate context in which they were conducted.

In conclusion, our findings challenge the notion that symptom status alone can be used to meaningfully stratify TB disease severity, and instead point to setting and diagnostic algorithm design as key determinants of the spectrum of disease that is identified. Both sTB and aTB encompass substantial heterogeneity, particularly when comparing people detected through clinic-based diagnosis versus active community screening. These results demonstrate the need for caution when interpreting and generalising findings across studies that use different diagnostic algorithms and settings, and for clear descriptions of how and where people are identified. Moving beyond symptom-based dichotomies toward a framework that integrates diagnostic context and objective measures of disease severity may provide a more accurate and useful understanding of the TB disease spectrum.

## Acknowledgements

Author contributions: NM, IG, ADG, and PYK conceived the idea for the manuscript. NM led the writing of the manuscript, with input from IG, ADG, and PYK. NM conducted the data analysis and prepared the figures, with input from PYK. ADG, PYK, IG, and NM obtained funding for the clinic study. All authors reviewed and edited the manuscript.

We would like to thank all the participants and staff involved in the clinic study and community survey. A list of staff involved in the two studies is provided in the supporting material.

## Data availability

An anonymised dataset and analysis code are available on the AHRI Data Repository https://data.ahri.org/index.php/catalog/1268 and on LSHTM Data Compass https://datacompass.lshtm.ac.uk/id/eprint/5244/

For the purposes of open access, the author has applied a Creative Commons Attribution (CC BY) licence to any Accepted Author Manuscript version arising from this submission.

