## Supporting material for "Influence of setting and diagnostic algorithm on disease severity among people diagnosed with symptomatic and asymptomatic tuberculosis in South Africa"

#### Additional methods

##### Community survey inclusion in analyses flowchart

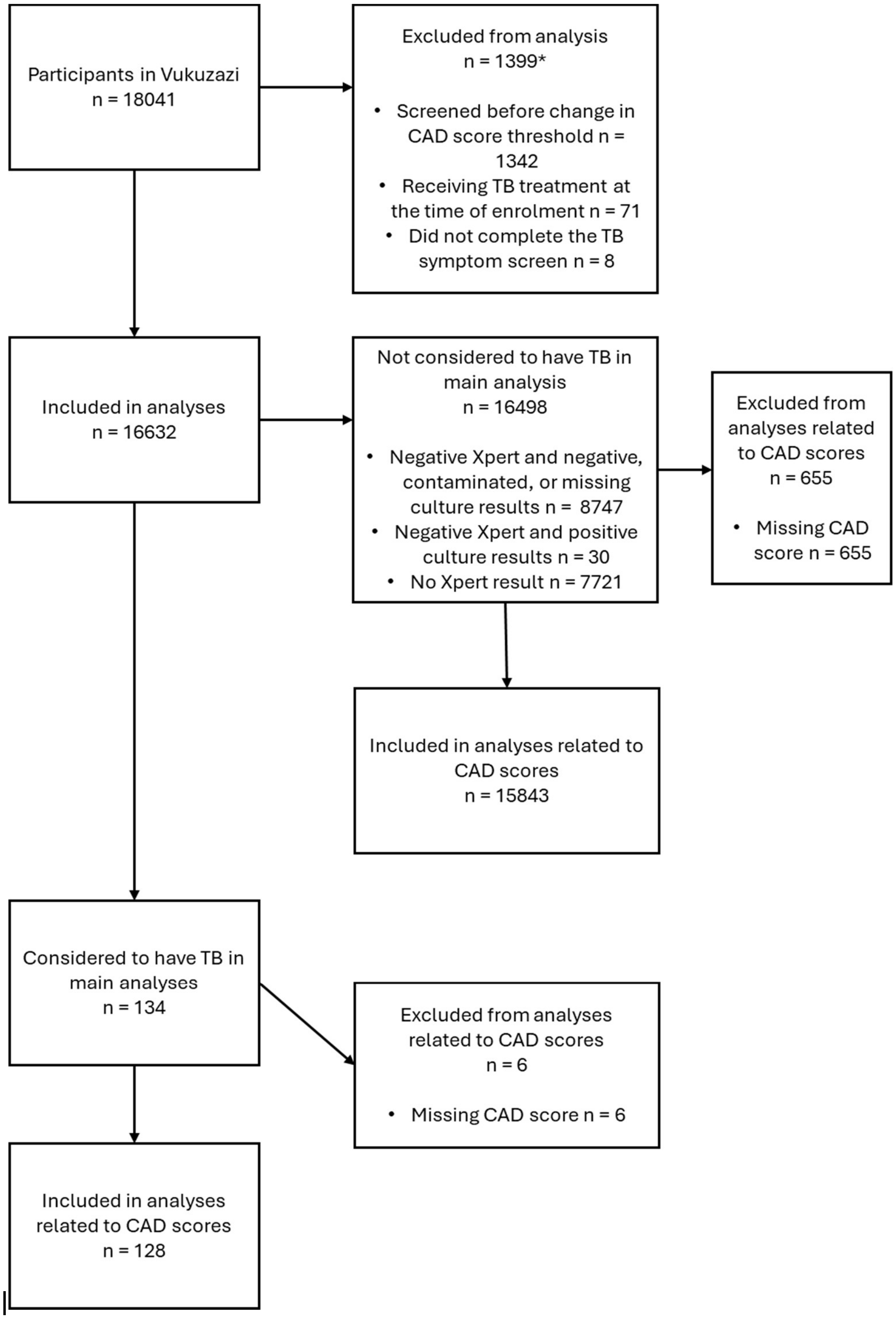


Figure S1. Flowchart showing the inclusion and exclusion of the community survey participants in the analyses

*Categories not mutually exclusive

#### Effect of choice of screening algorithms and CAD threshold

As the majority of people with CAD4TB score <25 who did not report symptoms were not tested, we estimated the proportion who would have been diagnosed with TB, and their CAD scores and Xpert result, had they been eligible for testing. This was done using bootstrapping, assuming that the prevalence of TB in people with CAD score <25 was the same for people who did not report symptoms as for people who did, and sampling CAD and Xpert scores for people with aTB and CAD score <25 from the CAD scores for people diagnosed with sTB with CAD score <25. As testing of people with CAD score<25 who did not reported symptoms was based on clinical indications, rather than conducted at random, we discarded the Xpert test results for those individuals in these analyses. To give an indication as to whether the assumptions underlying the bootstrapped estimates are likely to be reasonable, we compared the prevalence of TB, median CAD scores, and the proportion with Xpert result greater than trace between people with CAD scores of 25-49 diagnosed with aTB vs sTB.

### Additional results

#### Results including people in the community survey who were Xpert-undetected, culture-positive

| | | By setting | | | By setting and reported symptoms | | | | | |
|---|---|---|---|---|---|---|---|---|---|---|
| | | Community | Clinics | | Community | | | Clinics | | |
| | | | | Difference in proportion (95% CI) | Asymptomatic | Symptomatic | Difference in proportion (95% CI) | Asymptomatic | Symptomatic | Difference in proportion (95% CI) |
| Total (row percentages) | | 164 (31.2%) | 362 (68.8%) | | 138 (84.1%) | 26 (15.9%) | | 50 (13.8%) | 312 (86.2%) | 164 (31.2%) |
| | | **Col n (%)** | **Col n (%)** | | **Col n (%)** | **Col n (%)** | | **Col n (%)** | **Col n (%)** | |
| Sex | Male | 74 (45.1%) | 178 (49.2%) | -4.0% (-13.2%, 5.1%) | 60 (43.5%) | 14 (53.8%) | 10.4% (-10.5%, 31.2%) | 16 (32.0%) | 162 (51.9%) | 19.9% (5.9%, 34.0%) |
| | Female | 90 (54.9%) | 184 (50.8%) | 4.0% (-5.1%, 13.2%) | 78 (56.5%) | 12 (46.2%) | -10.4% (-31.2%, 10.5%) | 34 (68.0%) | 150 (48.1%) | -19.9% (-34.0%, -5.9%) |
| Age (years) | 15 - 29 | 37 (22.6%) | 102 (28.2%) | -5.6% (-13.5%, 2.3%) | 32 (23.2%) | 5 (19.2%) | -4.0% (-20.7%, 12.7%) | 16 (32.0%) | 86 (27.6%) | -4.4% (-18.3%, 9.4%) |
| | 30 - 44 | 43 (26.2%) | 149 (41.2%) | -14.9% (-23.4%, -6.5%) | 40 (29.0%) | 3 (11.5%) | -17.4% (-31.9%, -3.0%) | 19 (38.0%) | 130 (41.7%) | 3.7% (-10.9%, 18.2%) |
| | 45+ | 84 (51.2%) | 111 (30.7%) | 20.6% (11.6%, 29.6%) | 66 (47.8%) | 18 (69.2%) | 21.4% (1.8%, 41.0%) | 15 (30.0%) | 96 (30.8%) | 0.8% (-12.9%, 14.5%) |
| Past TB treatment | No | 130 (79.3%) | 275 (76.0%) | 3.3% (-4.3%, 10.9%) | 112 (81.2%) | 18 (69.2%) | -11.9% (-30.8%, 7.0%) | 39 (78.0%) | 236 (75.6%) | -2.4% (-14.8%, 10.1%) |
| | Yes | 34 (20.7%) | 87 (24.0%) | -3.3% (-10.9%, 4.3%) | 26 (18.8%) | 8 (30.8%) | 11.9% (-7.0%, 30.8%) | 11 (22.0%) | 76 (24.4%) | 2.4% (-10.1%, 14.8%) |
| BMI (kg/m2) | Underweight | 12 (7.36%) | 69 (19.1%) | -11.7% (-17.4%, -6.1%) | 8 (5.80%) | 4 (16.0%) | 9.6% (-4.8%, 24.0%) | 4 (8.00%) | 65 (20.8%) | 12.8% (4.1%, 21.6%) |
| | Normal | 86 (52.8%) | 185 (51.1%) | 1.3% (-7.9%, 10.6%) | 74 (53.6%) | 12 (48.0%) | -7.5% (-28.4%, 13.4%) | 19 (38.0%) | 166 (53.2%) | 15.2% (0.7%, 29.8%) |
| | Overweight/ obese | 65 (39.9%) | 108 (29.8%) | 9.8% (1.0%, 18.6%) | 56 (40.6%) | 9 (36.0%) | -6.0% (-26.0%, 14.1%) | 27 (54.0%) | 81 (26.0%) | -28.0% (-42.7%, -13.4%) |

| | | | | | | | | | | |
|---|---|---|---|---|---|---|---|---|---|---|
| HIV/ART | HIV- | 93 (57.8%) | 129 (35.6%) | 21.1% (12.0%, 30.1%) | 81 (60.0%) | 12 (46.2%) | -12.5% (-33.4%, 8.3%) | 11 (22.0%) | 118 (37.8%) | 15.8% (3.1%, 28.5%) |
| | HIV+ART- | 10 (6.21%) | 28 (7.73%) | -1.6% (-6.2%, 2.9%) | 9 (6.67%) | 1 (3.85%) | -2.7% (-11.1%, 5.8%) | 1 (2.00%) | 27 (8.65%) | 6.7% (1.7%, 11.6%) |
| | HIV+ART+ | 58 (36.0%) | 205 (56.6%) | -21.3% (-30.2%, -12.3%) | 45 (33.3%) | 13 (50.0%) | 17.4% (-3.4%, 38.1%) | 38 (76.0%) | 167 (53.5%) | -22.5% (-35.5%, -9.4%) |
| Cavitation | No | 124 (79.0%) | 214 (68.6%) | 10.4% (2.2%, 18.6%) | 105 (79.5%) | 19 (76.0%) | -3.5% (-21.6%, 14.6%) | 36 (94.7%) | 178 (65.0%) | -29.8% (-38.8%, -20.7%) |
| | Yes | 33 (21.0%) | 98 (31.4%) | -10.4% (-18.6%, -2.2%) | 27 (20.5%) | 6 (24.0%) | 3.5% (-14.6%, 21.6%) | 2 (5.26%) | 96 (35.0%) | 29.8% (20.7%, 38.8%) |
| CAD score (CAD4TB v5) | <50 | 48 (32.0%) | 95 (32.1%) | -0.1% (-9.3%, 9.1%) | 42 (33.1%) | 6 (26.1%) | -7.0% (-26.7%, 12.7%) | 18 (46.2%) | 77 (30.0%) | -16.2% (-32.8%, 0.4%) |
| | 50 - 69 | 51 (34.0%) | 44 (14.9%) | 19.1% (10.5%, 27.7%) | 41 (32.3%) | 10 (43.5%) | 11.2% (-10.6%, 33.0%) | 9 (23.1%) | 35 (13.6%) | -9.5% (-23.3%, 4.4%) |
| | 70 - 89 | 35 (23.3%) | 61 (20.6%) | 2.7% (-5.5%, 10.9%) | 30 (23.6%) | 5 (21.7%) | -1.9% (-20.3%, 16.5%) | 7 (17.9%) | 54 (21.0%) | 3.1% (-10.0%, 16.1%) |
| | 90 - 100 | 16 (10.7%) | 96 (32.4%) | -21.8% (-29.0%, -14.5%) | 14 (11.0%) | 2 (8.70%) | -2.3% (-15.1%, 10.4%) | 5 (12.8%) | 91 (35.4%) | 22.6% (10.6%, 34.6%) |
| Xpert | Undetected | 30 (18.3%) | 0 (0%) | 18.3% (12.4%, 24.2%) | 23 (16.7%) | 7 (26.9%) | 10.3% (-7.9%, 28.4%) | 0 (0%) | 0 (0%) | 0.0% (0.0%, 0.0%) |
| | Trace | 61 (37.2%) | 86 (23.8%) | 13.4% (4.8%, 22.0%) | 53 (38.4%) | 8 (30.8%) | -7.6% (-27.1%, 11.9%) | 22 (44.0%) | 64 (20.5%) | -23.5% (-38.0%, -9.0%) |
| | Greater than trace | 73 (44.5%) | 276 (76.2%) | -31.7% (-40.5%, -23.0%) | 62 (44.9%) | 11 (42.3%) | -2.6% (-23.3%, 18.1%) | 28 (56.0%) | 248 (79.5%) | 23.5% (9.0%, 38.0%) |
| Culture time to positivity | <10 days | 14 (8.81%) | 73 (21.6%) | -11.6% (-17.6%, -5.7%) | 11 (8.21%) | 3 (12.0%) | 3.6% (-9.5%, 16.7%) | 1 (2.04%) | 72 (24.9%) | 21.1% (15.0%, 27.2%) |
| | 10 - 15 days | 31 (19.5%) | 48 (14.2%) | 5.6% (-1.3%, 12.6%) | 24 (17.9%) | 7 (28.0%) | 9.5% (-8.7%, 27.7%) | 2 (4.08%) | 46 (15.9%) | 10.7% (4.0%, 17.5%) |
| | 15 - 42 days | 50 (31.4%) | 54 (16.0%) | 15.6% (7.6%, 23.5%) | 42 (31.3%) | 8 (32.0%) | 0.3% (-19.0%, 19.7%) | 6 (12.2%) | 48 (16.6%) | 3.4% (-6.5%, 13.2%) |
| | Culture negative | 64 (40.3%) | 163 (48.2%) | -6.0% (-15.1%, 3.1%) | 57 (42.5%) | 7 (28.0%) | -14.4% (-33.3%, 4.5%) | 40 (81.6%) | 123 (42.6%) | -40.6% (-52.9%, -28.2%) |

Table S2. Demographic and clinical characteristics of people diagnosed with aTB or aTB in the clinic and community surveys, including people with Xpert undetected, culture positive TB. In people diagnosed with TB as part of the community survey, data were missing on BMI for one individual, HIV/ART for three individuals, cavitation and CAD score for seven individuals, and time to culture positivity for five individuals. In people diagnosed with TB in clinics, data were missing on cavitation from 50 individuals, CAD score from 49 individuals and time to culture positivity for 24 individuals. Confidence intervals for differences in proportions (risk difference) are estimated using the score method.

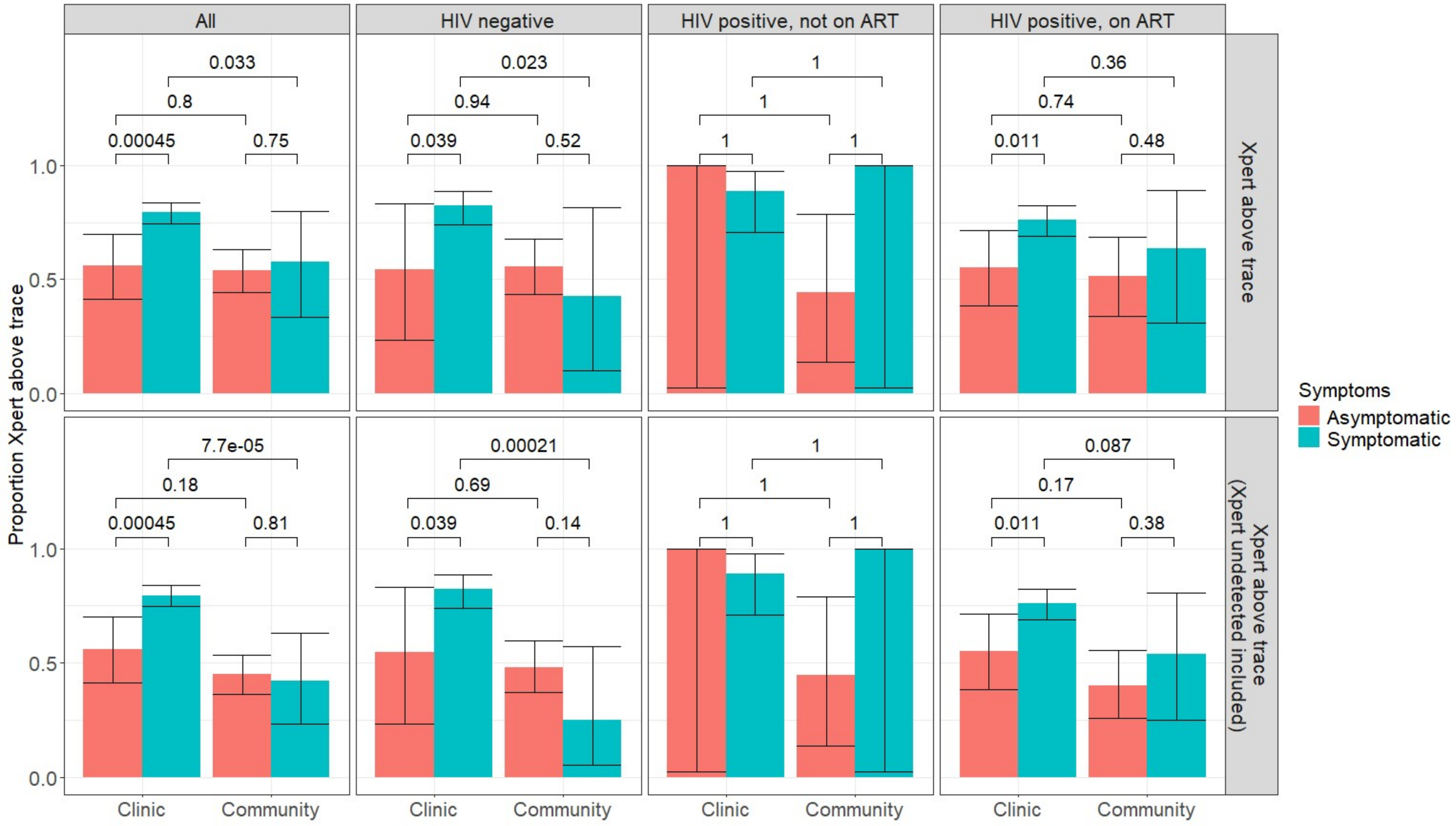


Figure S2. Comparison of the proportion of people who were Xpert trace or above in people diagnosed with TB, by setting (clinic vs community), symptoms, and HIV/ART status. People detected in the community survey who were Xpert undetected and culture positive were excluded from the analysis in the top row, and included in the analysis in the bottom row. Error bars show 95% confidence intervals, and the numbers on the plots are p-values, calculated using a Wald test.

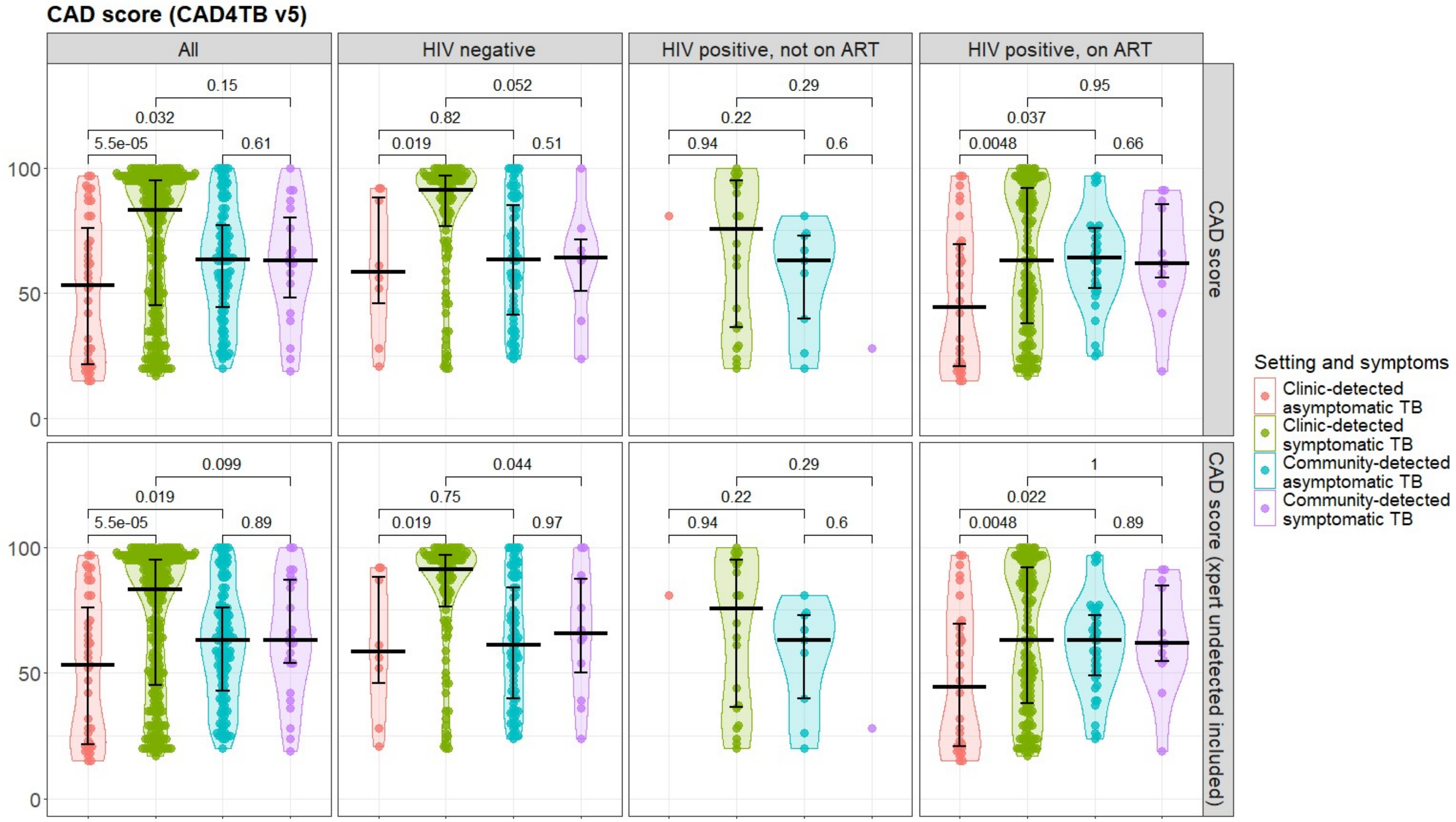


Figure S3. Comparison of mean CAD scores in people diagnosed with TB, by setting (clinic vs community), symptoms, and HIV/ART status. People detected in the community survey who were Xpert undetected and culture positive were excluded from the analysis in the top row, and included in the analysis in the bottom row. Horizontal lines show median and interquartile range, and the numbers on the plots are p-values, calculated using a Mann-Whitney U-Test

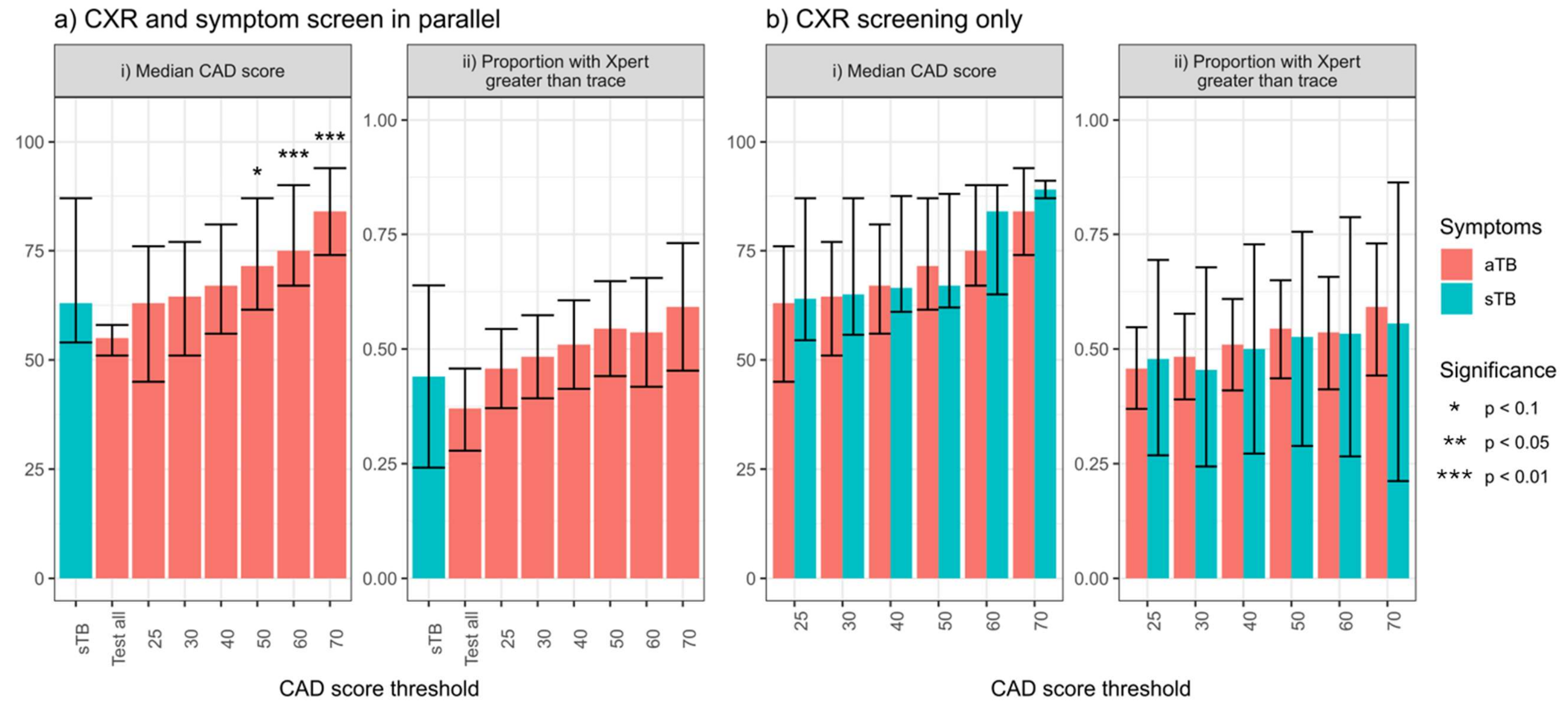


Figure S4. Effect of choice of CAD4TB v5 cut-off used in screening on relative severity of aTB and sTB diagnosed in the community, including people diagnosed with Xpert undetected, culture positive TB.

In a), it assumed that all people who report symptoms will be eligible for testing, but people who do not report symptoms will be eligible for testing only if they have a CAD score above the threshold. In b) it is assumed that people will be eligible for testing only if they have a CAD score above the threshold, regardless of reported symptoms. The bars Plots a – d: Asterisks indicate p-values for differences between aTB and sTB, where $p<0.1$. Where no asterisk is shown, $p\geq0.1$. i) The bars show the median, and the error bars the inter-quartile range. p-values calculated using a Mann-Whitney U-Test. ii) The bars show the mean, and the error bars the 95% confidence interval. p-values calculated using a Wald test

#### Estimated prevalence and severity of Xpert Ultra positive TB by CAD score and reported symptoms

Table S2 shows that there was little difference in the estimated prevalence or severity of TB disease between people with CAD scores of 25 – 49 who did or did not report TB symptoms, providing support for the assumptions made in estimating TB prevalence and severity in people with CAD score <25 who did not report symptoms.

| CAD4TB v5 score | Total | | Xpert Ultra positive TB (prevalence/100,000) | | Median CAD score | | Proportion with Xpert Ultra result greater than trace | |
|---|---|---|---|---|---|---|---|---|
| | No reported symptoms | Reported symptoms | No reported symptoms | Reported symptoms | No reported symptoms | Reported symptoms | No reported symptoms | Reported symptoms |
| 0 – 24 | 6,754 | 407 | 2 (30) + | 2 (489) | 22 | 21.5 | 1.00 | 0.00 |
| 25 – 49 | 5,931 | 547 | 30 (506) | 3 (545) | 35 | 39 | 0.35 | 0.33 |
| 50 – 74 | 1,654 | 235 | 44 (2,660) | 8 (3,292) | 63.5 | 63 | 0.65 | 0.67 |
| 75 + | 347 | 77 | 33 (9,510) | 6 (7,229) | 89.5 | 89 | 0.65 | 0.86 |
| Missing* | 605 | 46 | 6 (992) | 0 | | | 0.83 | |
| Total | 15,291 | 1,312 | 115 (752) | 19 (1,427) | 64 | 64 | 0.55 | 0.62 |

Table S1. Estimated TB prevalence and severity (median CAD scores and proportion with Xpert Ultra result greater than trace) in the community survey, by reported symptoms and CAD score

+People with CAD score <25 were only tested if they reported TB symptoms or if testing was indicators for other reasons (e.g. other reported symptoms), and so figures for CAD score 0 – 24 likely underestimate the prevalence of TB in people who did not report symptoms

*10 people with missing CAD score had missing data on reports symptoms, and are not included in the table

#### Acknowledgements

**Vukuzazi Team** - Below is a list of staff that significantly contributed to the implementation and conduct of the community study.

| Name | Role |
|---|---|
| Deenan Pillay | Principal Investigator (2017-2019) |
| Willem Hanekom | Principal Investigator (2019-present) |
| Emily Wong | Co-Principal Investigator |
| Mark Siedner | Co-Principal Investigator |
| Olivier Koole | Co-Principal Investigator (2017-2019) |
| Thumbi Ndung'u | Co-investigator |
| Thandeka Khoza | Co-investigator (2019-present) |
| Kobus Herbst | Co-investigator |
| Kathy Baisley | Co-investigator |
| Janet Seeley | Co-investigator |
| Alison Grant | Co-investigator |
| Resign Gunda | Programme Manager |
| Ashmika Surujdeen | Study Coordinator |
| Theresa Smit | Head: Diagnostic Research |
| Dickman Gareta | Head: Research Data Management |
| Day Munatsi | Head: Research Data Systems |
| Ngcebo Mhlongo | Study Physician |
| Tshwaraganang Modise | Research Data Manager |
| Jaco Dreyer | Senior Research Data Manager |
| Siyabonga Nxumalo | Research Data Manager |
| Stephen Olivier | Statistician |
| Gregory Ording-Jespersen | Laboratory Data Supervisor |
| Innocentia Mpofana | Diagnostic Laboratory Manager |
| Khadija Khan | Biorepository Manager |
| Zizile Sikhosana | Somkhele Laboratory Supervisor |
| Sashen Moodley | Microbiology Laboratory Supervisor |
| Hollis Shen | Head: Exploratory Research Division |
| Philippa Mathews | Clinical Governance |
| Nompilo Buthelezi | Training Coordinator |
| Hlolisile Khumalo | Nursing Manager |
| Sanah Bucibo | Professional Nurse |
| Nozipho Mbonambi | Professional Nurse |
| Hloniphile Ngubane | Professional Nurse |
| Thokozani Simelane | Professional Nurse |
| Khanyisani Buthelezi | Professional Nurse |
| Sphiwe Ntuli | Professional Nurse |
| Nombuyiselo Zondi | Professional Nurse |
| Siboniso Nene | Professional Nurse |
| Bongumenzi Ndlovu | Enrolled Nurse |
| Talente Ntimbane | Enrolled Nurse |

| | |
|---|---|
| Mbali Mbuyisa | Enrolled Nurse |
| Xolani Mkhize | Enrolled Nurse |
| Melusi Sibiya | Enrolled Nurse |
| Ntombiyenkosi Ntombela | Enrolled Nurse |
| Mandisi Dlamini | Enrolled Nurse |
| Hlobisile Chonco | Enrolled Nurse |
| Hlengiwe Dlamini | Enrolled Nurse |
| Doctar Mlambo | Enrolled Nurse |
| Nonhlanhla Mzimela | Enrolled Nurse |
| Zinhle Buthelezi | Enrolled Nurse |
| Zinhle Mthembu | Enrolled Nurse |
| Thokozani Bhengu | Enrolled Nurse |
| Sandile Mthembu | Enrolled Nurse |
| Phumelele Mthethwa | Enrolled Nurse |
| Zamashandu Mbatha | Enrolled Nurse |
| Welcome Petros Mthembu | Enrolled Nurse |
| Anele Mkhwanazi | Clinical Research Assistant Supervisor |
| Mandlakayise Zikhali | Clinical Research Assistant Supervisor |
| Phakamani Mkhwanazi | Clinical Research Assistant |
| Ntombiyenhlanhla Mkhwanazi | Clinical Research Assistant |
| Rose Myeni | Clinical Research Assistant |
| Fezeka Mfeka | Clinical Research Assistant |
| Hlobisile Gumede | Clinical Research Assistant |
| Nonceba Mfeka | Clinical Research Assistant |
| Ayanda Zungu | Clinical Research Assistant |
| Nonhlanhla Mfekayi | Clinical Research Assistant |
| Smangaliso Zulu | Clinical Research Assistant |
| Mzamo Buthelezi | Clinical Research Assistant |
| Senzeni Mkhwanazi | Clinical Research Assistant |
| Mlungisi Dube | Clinical Research Assistant |
| Hosea Kambonde | IT Systems Developer |
| Lindani Mthembu | Information Technology Assistant |
| Seneme Mchunu | Information Technology Assistant |
| Sibahle Gumbi | Research Admin Assistant |
| Tumi Madolo | Research Data Manager |
| Thengokwakhe Nkosi | Driver |
| Sibusiso Mkhwanazi | Driver |
| Sibusiso Nsibande | Driver |
| Mpumelelo Steto | Driver |
| Sibusiso Mhlongo | Driver |
| Velile Vellem | Driver |
| Pfarelo Tshivase | Driver |
| Jabu Kwinda | Driver |
| Bongani Magwaza | General Worker |
| Siyabonga Nsibande | General Worker |
| Skhumbuzo Mthombeni | General Worker |
| Sphiwe Clement Mthembu | General Worker |

| | |
|---|---|
| Antony Rapulana | Laboratory Technologist |
| Jade Cousins | Laboratory Technologist |
| Thabile Zondi | Laboratory Technologist |
| Nagavelli Padayachi | Laboratory Technologist |
| Freddy Mabetlela | Laboratory Technologist |
| Simphiwe Ntshangase | Laboratory Technician/LIMS Administrator |
| Nomfundo Luthuli | Laboratory Technician |
| Sithembile Ngcobo | Laboratory Technologist |
| Kayleen Brien | Laboratory Technologist |
| Sizwe Ndlela | Laboratory Technician |
| Nomfundo Ngema | Laboratory Technician |
| Nokukhanya Ntshakala | Laboratory Technician |
| Anupa Singh | Laboratory Technician |
| Rochelle Singh | Laboratory Technician |
| Logan Pillay | Laboratory Technician |
| Kandaseelan Chetty | Laboratory Technician |
| Ashentha Govender | Laboratory Technician |
| Pamela Ramkalawon | Laboratory Research Technician |
| Nondumiso Mabaso | Laboratory Intern |
| Kimeshree Perumal | Laboratory Intern |
| Senamile Makhari | Biorepository Laboratory Technician |
| Nondumiso Khuluse | Biorepository Laboratory Technician |
| Nondumiso Zitha | Biorepository Research Assistant |
| Hlengiwe Khathi | Biorepository Research Assistant |
| Mbuti Mofokeng | Clinical Specimen Driver/Laboratory Assistant |
| Nomathamsanqa Majozi | Public Engagement |
| Nceba Gqaleni | Public Engagement |
| Hannah Keal | Communications |
| Phumla Ngcobo | Communications |
| Costa Criticos | Operational Oversight |
| Raynold Zondo | Operational Oversight |
| Dilip Kalyan | Operational Oversight |
| Clive Mavimbela | Operational Oversight |
| Anand Ramnanan | Procurement |
| Sashin Harilall | Grants Office |
| Kennedy Nyamande | Pulmonology Consultant |
| Jaikrishna Kalideen | Radiologist |
| Ramesh Jackpersad | Radiologist |
| Kgaugelo Moropane | Radiographer |
| Boitsholo Mfolo | Radiographer |
| Khabonina Malomane | Radiographer |

#### AHCoS team

Many AHRI staff contributed to the clinic-based study, including Mzamo Buthelezi, Nqubelo Dladla, Mlungisi Dube, Nokuthula Jiyane, Nomthandazo Mbatha, Nonkululeko Magagula, Ginger Mahlake, Sphephelo Masondo, Pinkie Mbotho, Sanele Mdletshe, Edwin Mkhwanazi, Mazani Mkhwanazi, Sanele Mngadi, Silindile Mthembu, Sizwe Ndlela, Nokuthula P. Ndlovu, Mbuso Ngema, Nomthunzi N. Ngema, Nothando Nkosi, Mpepho S. Ntombela, Simphiwe B. Ntshangase, Siyabonga Nxumalo, Thuthukani Nyawo, Nolwazi Nyuswa, Bongekile Nzuza, Bathandekile M. Phahlamohlaka, Zizile E. Sikhosana, Sibongokuhle M. Zulu and Professor Rodney Ehrlich
